# Olfactory Receptor Expression Profiles in Clear Cell Renal Cell Carcinoma Reveal Potential Diagnostic and Prognostic Markers

**DOI:** 10.1101/2025.01.05.25320001

**Authors:** Kazuya Hasegawa, Yuya Yamaguchi

## Abstract

**Background:** Olfactory receptors (ORs) are G protein-coupled receptors that are aberrantly expressed in various cancers, including clear cell renal cell carcinoma (KIRC). However, the roles of ORs in KIRC are unknown. This study aimed to comprehensively analyze the expression profiles of OR genes in KIRC and evaluate their potential as diagnostic and prognostic biomarkers.

**Methods:** We analyzed RNA-seq data from The Cancer Genome Atlas KIRC dataset, which contains 72 normal and 530 tumor samples. We selected OR genes with median transcripts per million (TPM) values of 1 or higher in at least one group (normal or tumor). Differential expression analysis was performed using the Student’s t-test or Wilcoxon rank-sum test. The diagnostic potential of OR genes was evaluated using receiver operating characteristic (ROC) curve analysis. Kaplan-Meier analysis was employed to assess the association between OR gene expression and patient survival. Sex-based differences in OR gene expression were also examined.

**Results:** We identified 11 OR genes with significant changes in KIRC expression. Among them, OR2A4, OR51E1, and OR7E14P showed high diagnostic performance, with AUC values of 0.951, 0.924, and 0.910, respectively. Combining these three genes improved the AUC to 0.972. High OR2A20P expression was significantly associated with poorer prognosis, whereas high OR7E7P expression was associated with better prognosis. We also found sex-based differences in the expression of OR2A7, OR2I1P, and OR7E14P, with females exhibiting significantly higher expression.

**Conclusions:** Our findings suggest that ORs, especially OR2A4, OR51E1, and OR7E14P, could serve as potential diagnostic markers for KIRC. OR2A20P and OR7E7P may represent promising prognostic markers. The observed sex-based differences in OR gene expression highlight the need for personalized treatment of KIRC. Further studies are warranted to validate these findings and elucidate the functional roles of ORs in the pathogenesis of KIRC.

## Introduction

Kidney cancer is a cancer with high morbidity and mortality rates worldwide [1]. In 2020, approximately 430,000 new cases were diagnosed worldwide, and approximately 180,000 people died of the disease [2]. Recent data from China and the United States also indicate a significant burden, with over 77,000 new cases and 46,000 deaths estimated in China and over 71,000 new cases and 15,000 deaths in the United States in 2022 [3]. Renal cell carcinoma (RCC) is the most common histological type, accounting for approximately 90% of kidney cancer cases [4]. Furthermore, clear cell RCC (ccRCC, commonly abbreviated as KIRC) accounts for approximately 70-80% of all RCC cases and is the most common histological type [5, 6]. KIRC is characterized by heterogeneous histology and diverse genetic alterations, including mutations in VHL, PBRM1, and SETD2 genes [7, 8]. The Cancer Genome Atlas (TCGA) Research Network comprehensively characterized the molecular features of ccRCC, providing a valuable resource for understanding the disease [9]. In this study, we focused on KIRC. Smoking, obesity, hypertension, and genetic factors have been reported as factors that cause KIRC [10]. The 5-year survival rate of KIRC is relatively good for early-stage localized cancer, but it remains low for advanced and metastatic cancer. The 5-year survival rate for stage I is about 80-90%, whereas that for stage IV it drops to about 10-20% [11]. In recent years, with the advancement of treatments, such as molecular targeted drugs and immune checkpoint inhibitors, the prognosis of advanced KIRC has improved [12, 13], but challenges, such as difficulty in early detection, metastasis, recurrence, and drug resistance, remain. Therefore, it is an important clinical challenge to better understand the biological characteristics of kidney cancer and identify highly sensitive and accurate diagnostic markers, prognostic predictive markers, new biomarkers that contribute to personalized medicine, and new therapeutic targets.

Olfactory receptors (ORs) are membrane proteins that belong to the G protein-coupled receptor (GPCR) superfamily. There are approximately 400 functional OR genes and approximately 450 pseudogenes in humans [14, 15]. Pseudogenes are genes that arise during evolution through gene duplication but have lost their function because of the accumulation of mutations and are believed to not code for proteins [16]. The OR is mainly responsible for the reception of odorants in the olfactory epithelium, but recent studies have revealed that the OR is expressed in various tissues other than olfactory tissues. Furthermore, ORs in non-olfactory tissues are involved in various cellular responses, such as cell proliferation, apoptosis, and migration [17–19].

In recent years, abnormal expression of OR genes has been reported in many types of cancer, suggesting their involvement in tumor development, progression, metastasis, and drug sensitivity. It has also been reported that OR expression is associated with prognosis in some carcinomas, and attention has been focused on the possibility that ORs can be diagnostic and prognostic markers. However, few studies have investigated the role of ORs in kidney cancer, and their expression profiles and clinical significance are largely unknown.

In this study, we comprehensively analyzed the expression of OR genes in kidney cancer using the KIRC dataset of The Cancer Genome Atlas (TCGA), a large-scale cancer genome dataset. We hypothesized that specific OR genes are specifically expressed in kidney cancer and that they are useful as diagnostic, prognostic, and therapeutic markers. To test this hypothesis, we compared the expression of OR genes in normal and tumor tissues and identified OR genes whose expression was significantly altered in tumor tissues. We then used receiver operating characteristic (ROC) analysis to evaluate the ability to discriminate between tumor and normal tissues and further used Kaplan-Meier analysis to examine the association between OR gene expression and patient survival. We also investigated gender differences in OR gene expression in tumor tissues. This study aimed to elucidate the role of OR genes in kidney cancer, which may contribute to the early detection, prognosis prediction, and personalized medicine of kidney cancer.

## Methods

### Data Source and Patient Cohort

Gene expression data and clinical information were obtained from The Cancer Genome Atlas (TCGA) Kidney Renal Clear Cell Carcinoma (KIRC) dataset (data release 41.0 - August 28, 2023). The dataset included 72 normal and 543 tumor samples. Tumor samples with duplicated RNA-seq data from the same patient were resolved by selecting the sample with the highest overall expression value. Samples without stage information were excluded from the analysis. The final tumor cohort consisted of 527 samples, with the following stage distribution: Stage I (n=265), Stage II (n=56), Stage III (n=123), and Stage IV (n=83).

### Preprocessing Gene Expression Data

RNA-seq data were quantified as transcripts per million (TPM) from TCGA. We transformed raw TPM values to log2(TPM+1) to reduce skewness in the RNA-seq data. For each sample, we plotted a box plot of the log2-transformed expression values for all genes. Genes with TPM < 1 in all samples were excluded to eliminate noise. We specifically focused on olfactory receptor (OR) genes, including those annotated as pseudogenes. To select OR genes with potentially relevant expression, we calculated the median TPM value for each OR gene in normal samples and each tumor stage (Stage I, II, III, and IV). Only OR genes with a median TPM value of 1 or higher in at least one of the five groups were included in subsequent analyses.

### Differential Expression Analysis

Differential expression analysis between normal and tumor samples was performed using the Mann– Whitney U test. The Benjamini-Hochberg method was used to control the false discovery rate (FDR) and correct for multiple hypothesis testing.

### Receiver Operating Characteristic (ROC) curve analysis

To evaluate the diagnostic potential of OR genes, we performed receiver operating characteristic (ROC) curve analysis. The area under the receiver operating characteristic curve (AUC) was calculated for each OR gene to assess its ability to discriminate between tumor and normal samples.

### Survival Analysis

Overall survival time was defined as the time from initial diagnosis to death (days_to_death) for deceased patients and to the last follow-up (days_to_last_follow_up) for alive patients. Patients with a survival time of 29 days were excluded from the survival analysis to ensure sufficient observation time for all included cases. Patients who remained alive were censored as alive. To assess the prognostic value of OR genes, we performed Kaplan-Meier survival analysis. Tumor samples were divided into high and low expression groups based on the median log2(TPM+1) value of each OR gene in the tumor group. Samples with expressions equal to the median were assigned to the high-expression group. Survival curves were generated using the Kaplan-Meier method, and the log-rank test was used to compare survival distributions between high and low expression groups.

### Sex-Based Expression Analysis

To investigate potential sex-specific differences in OR gene expression, we compared expression levels between male (n=340) and female (n=187) patients within the tumor cohort using the two-sample t-test.

### Statistical Analysis

All statistical analyses were performed using JMP Pro 17 (SAS Institute Inc., Cary, NC, USA) and GraphPad Prism 10 (GraphPad Software, San Diego, CA, USA). JMP Pro 17 was used for the box plots. For the t-tests and ANOVA, GraphPad Prism 10 was used. ROC analysis and Kaplan-Meier survival analysis were conducted using JMP Pro 17. Statistical significance was defined as a two-sided P-value < 0.05.

## Results

### Altered Expression of Olfactory Receptor Genes in Renal Cell Carcinoma

To investigate the expression profiles of olfactory receptor (OR) genes in kidney cancer, we analyzed RNA-seq data from 72 normal kidney tissue samples and 527 kidney renal clear cell carcinoma (KIRC) tumor samples from The Cancer Genome Atlas (TCGA) KIRC dataset. We focused on OR genes that showed a median transcripts per million (TPM) value of 1 or higher in either the normal group or any of the tumor stage groups (Stage I, II, III, or IV), or both. This resulted in the identification of 11 OR genes (OR2A1-AS1, OR2A4, OR2A7, OR2A20P, OR2I1P, OR2T10, OR51E1, OR51E2, OR7E7P, OR7E14P, and OR7E47P).

First, to assess the overall trend of OR gene expression among the normal and different tumor stage groups, we summed the log2(TPM+1) values of the 11 OR genes for each sample and generated box plots for visualization (Figure 1). No statistically significant difference was observed in the sum of OR gene expression between the normal group and any of the tumor stage groups (Stage I-IV) (Figure 1). However, it is important to note that this summed expression includes genes with large differences in expression between the normal and tumor groups, and therefore, it requires careful interpretation.

**Figure 1.**
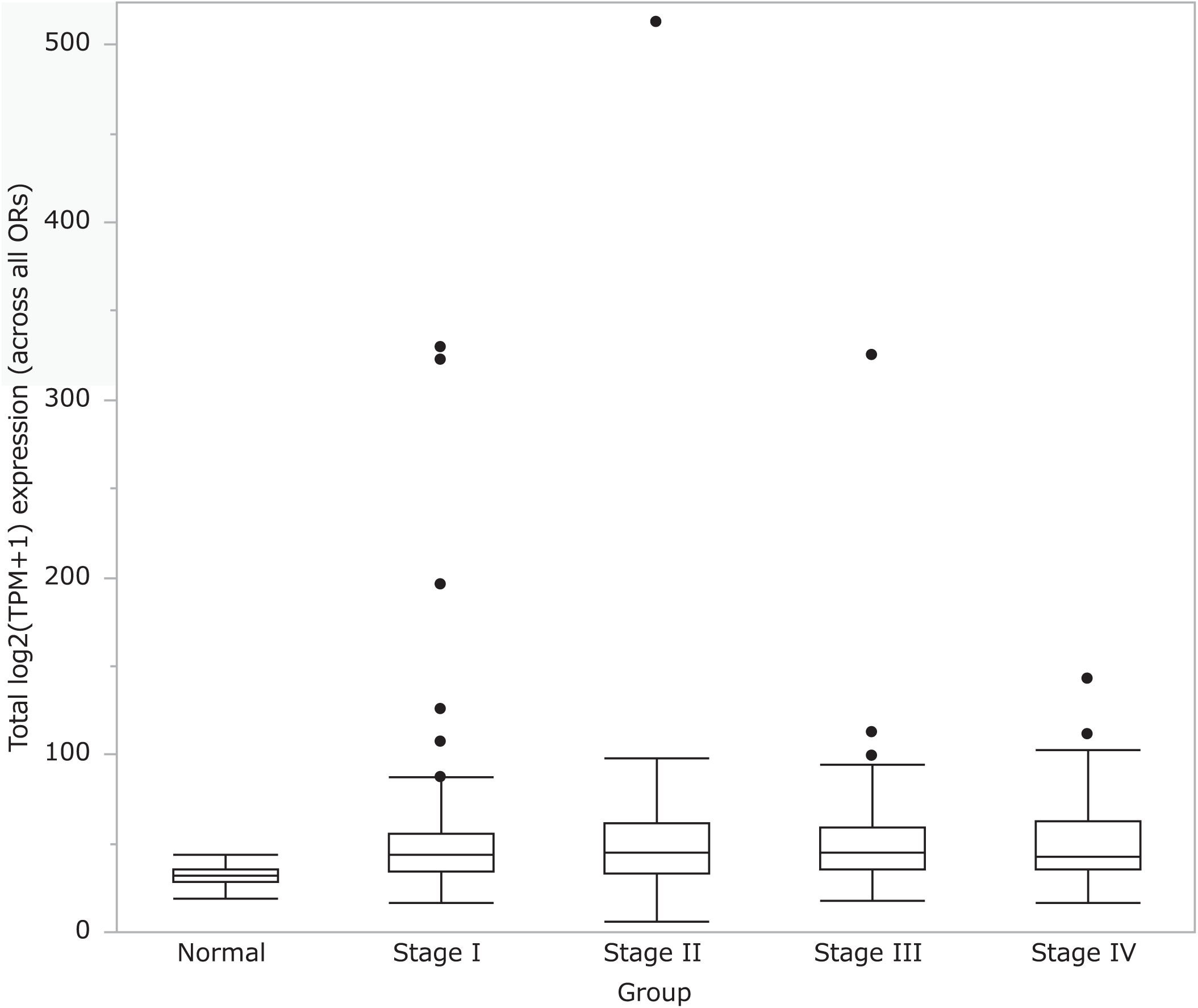
Box plot of the total log2(TPM+1) expression per sample (summed across 11 OR genes) in the normal group and each tumor stage group (Stage I, II, III, and IV). The boxes represent the interquartile range (IQR), the horizontal lines within the boxes denote the median, and the whiskers extend to 1.5 × IQR. Dots beyond whiskers indicate statistical outliers. No significant difference was found between the normal group and any stage group using one-way ANOVA.

Next, we compared the median expression patterns of the 11 OR genes between the normal group and each stage (Stage I to IV) of the tumor group, as shown in Figure 2. The heatmap displays the median log2(TPM+1) expression levels of each gene, with samples categorized into the normal group and tumor stages I-IV, and genes represented by the 11 selected OR genes. The heatmap reveals two distinct expression patterns. One group of OR genes (OR2A4, OR51E1, OR51E2, OR7E47P, OR2I1P, and OR7E7P) generally showed relatively higher median expression in the tumor groups compared with the normal group, although OR7E47P exhibited a slight decrease in expression in Stage IV. Another group of OR genes (OR2A1-AS1, OR2A20P, OR2A7, OR7E14P, and OR2T10) tended to exhibit lower median expression in the tumor groups than in the normal group.

**Figure 2.**
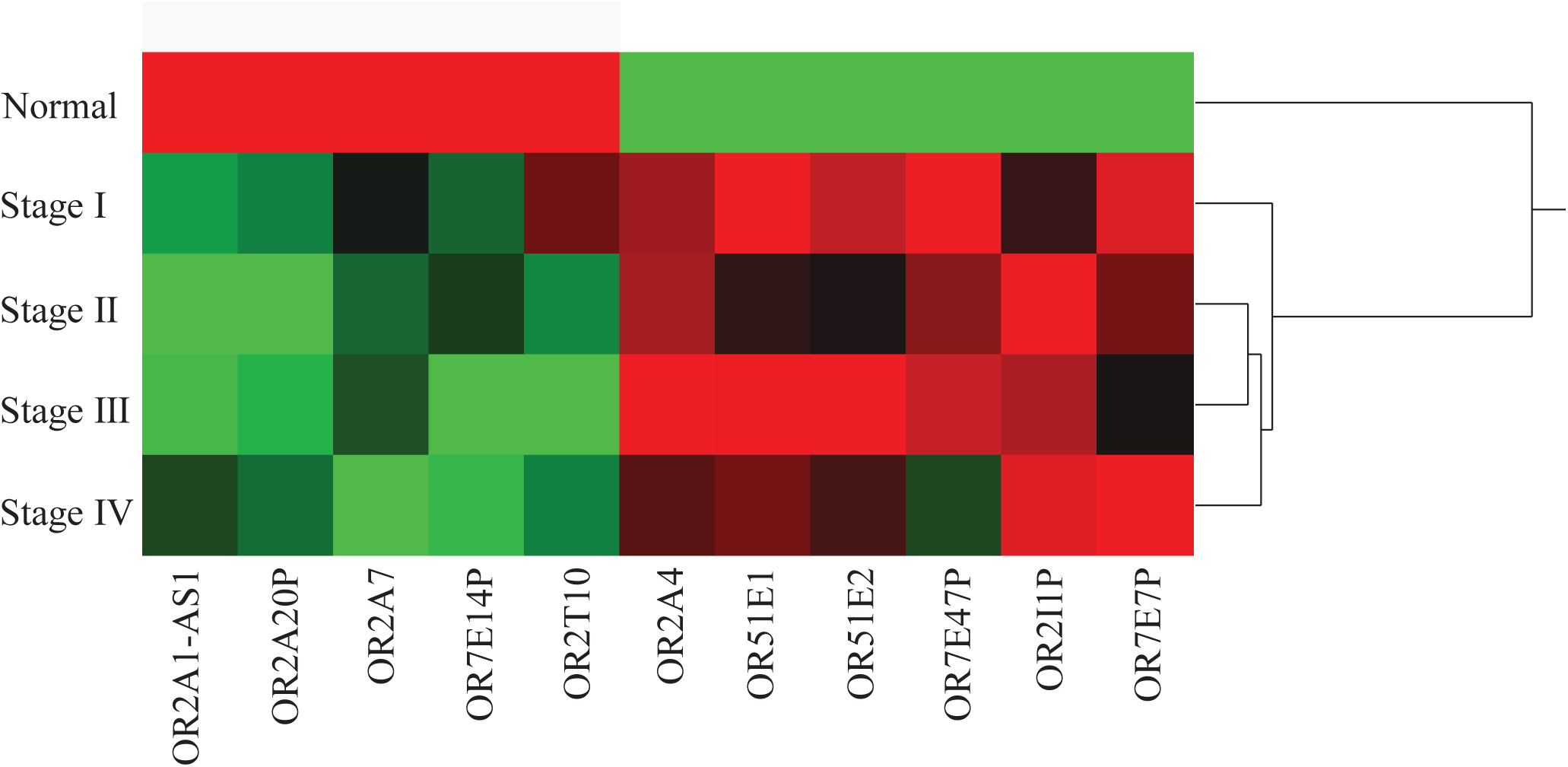
Heatmap showing the median log2(TPM+1) expression levels of 11 selected OR genes in normal kidney and KIRC samples (stages I-IV). This heatmap displays the median log2(TPM+1) expression levels of the 11 selected olfactory receptor (OR) genes across normal kidney (Normal) and KIRC samples (Stage I to Stage IV). Each row corresponds to a specific OR gene, with colors ranging from green (relatively low expression) to red (relatively high expression), indicating the median expression level across the designated groups. Hierarchical clustering was applied to identify expression pattern similarities among genes.

Furthermore, individual comparison of the expression levels of the 11 OR genes between the normal and tumor groups revealed that 10 OR genes showed significant changes in expression in the tumor group (p < 0.05, Figure 3). Specifically, six genes (OR2A4, OR51E1, OR51E2, OR7E47P, OR7E7P, and OR2I1P) were significantly upregulated in the tumor group, whereas four genes (OR2A1-AS1, OR2A7, OR7E14P, and OR2A20P) were significantly downregulated (Figure 3).

**Figure 3.**
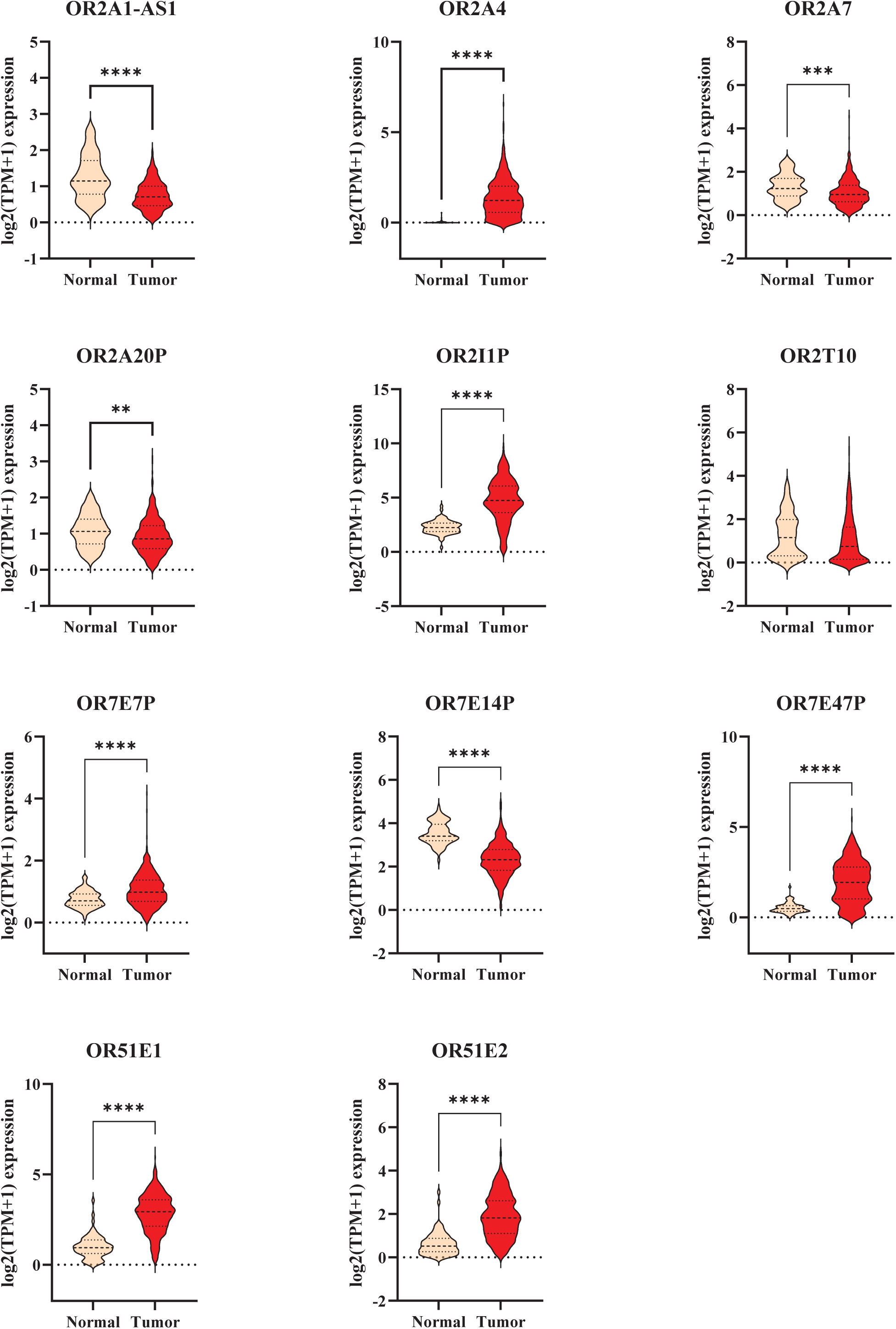
Comparison of OR gene expression between the normal and tumor groups. Box plots showing the expression levels of 11 OR genes in normal (n=72) and tumor (n=530) samples. *p < 0.05, **p < 0.01, ***p < 0.001, ****p < 0.0001 (Mann-Whitney U test).

When comparing OR gene expression across different tumor stages, significant differences were observed in only one gene (OR7E47P) (Figure 4). However, this gene did not show a clear stage-dependent pattern, with expression levels consistently increasing or decreasing with the advancing stage.

**Figure 4.**
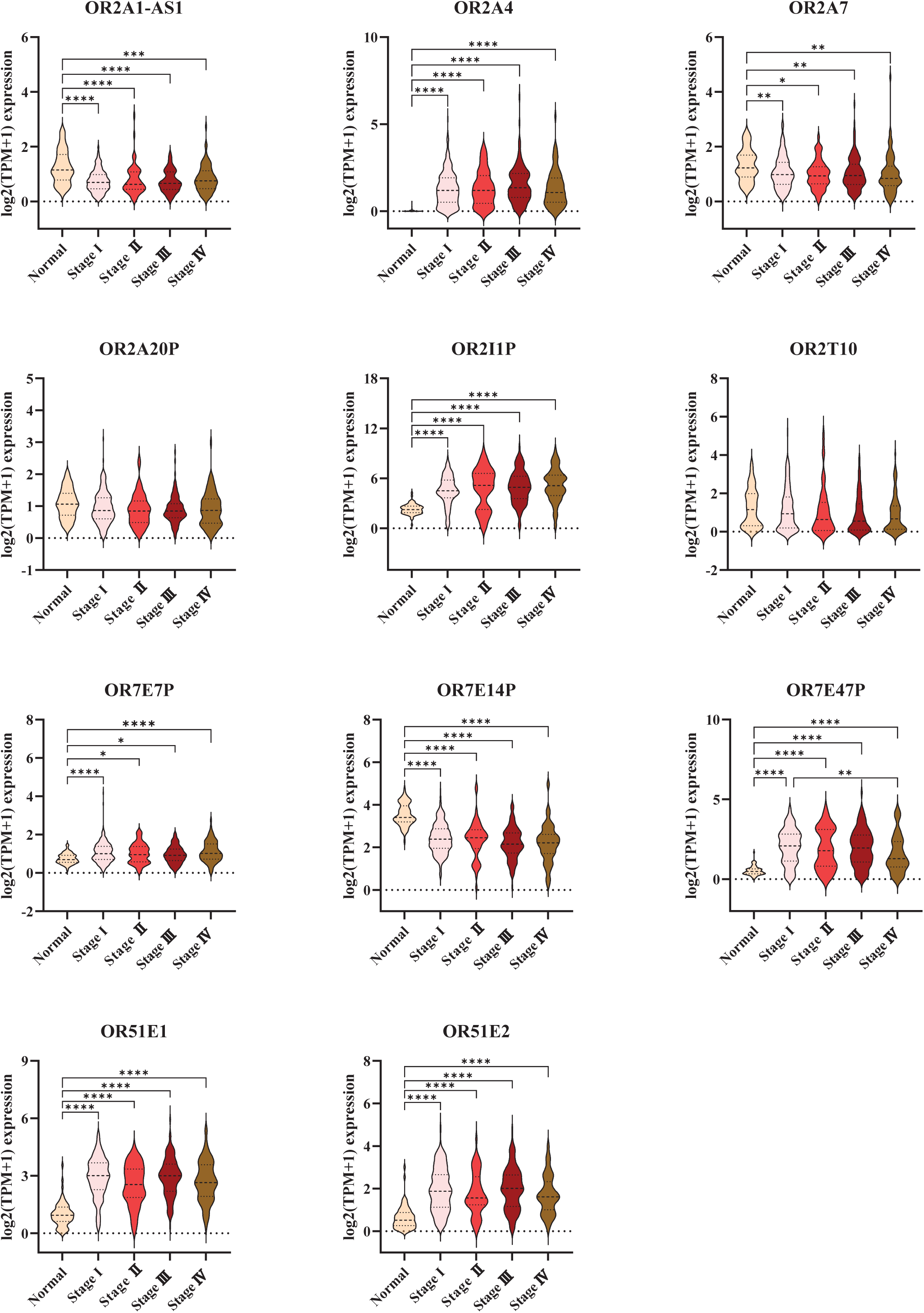
Comparison of OR gene expression among different tumor stages. Violin plots showing the expression of OR genes across different tumor stages (I-IV). A significant difference among the stages was found in only one gene (OR7E47P) using the Kruskal-Wallis test (*p < 0.05, **p < 0.01, ***p < 0.001, ****p < 0.0001).

### Potential of OR Genes as Diagnostic Markers

To evaluate the potential of OR genes as diagnostic markers for kidney cancer, we performed ROC curve analysis. First, we generated ROC curves for individual genes and assessed their ability to discriminate between normal and tumor samples. We found that OR2A4, OR51E1, and OR7E14P exhibited high AUC values even when considered alone (AUC = 0.951, 0.924, and 0.910, respectively, Figure 5). Furthermore, when these three genes were combined, the AUC reached 0.972, indicating excellent discriminatory ability (Figure 5). These results suggest that OR2A4, OR51E1, and OR7E14P are useful diagnostic markers for kidney cancer.

**Figure 5.**
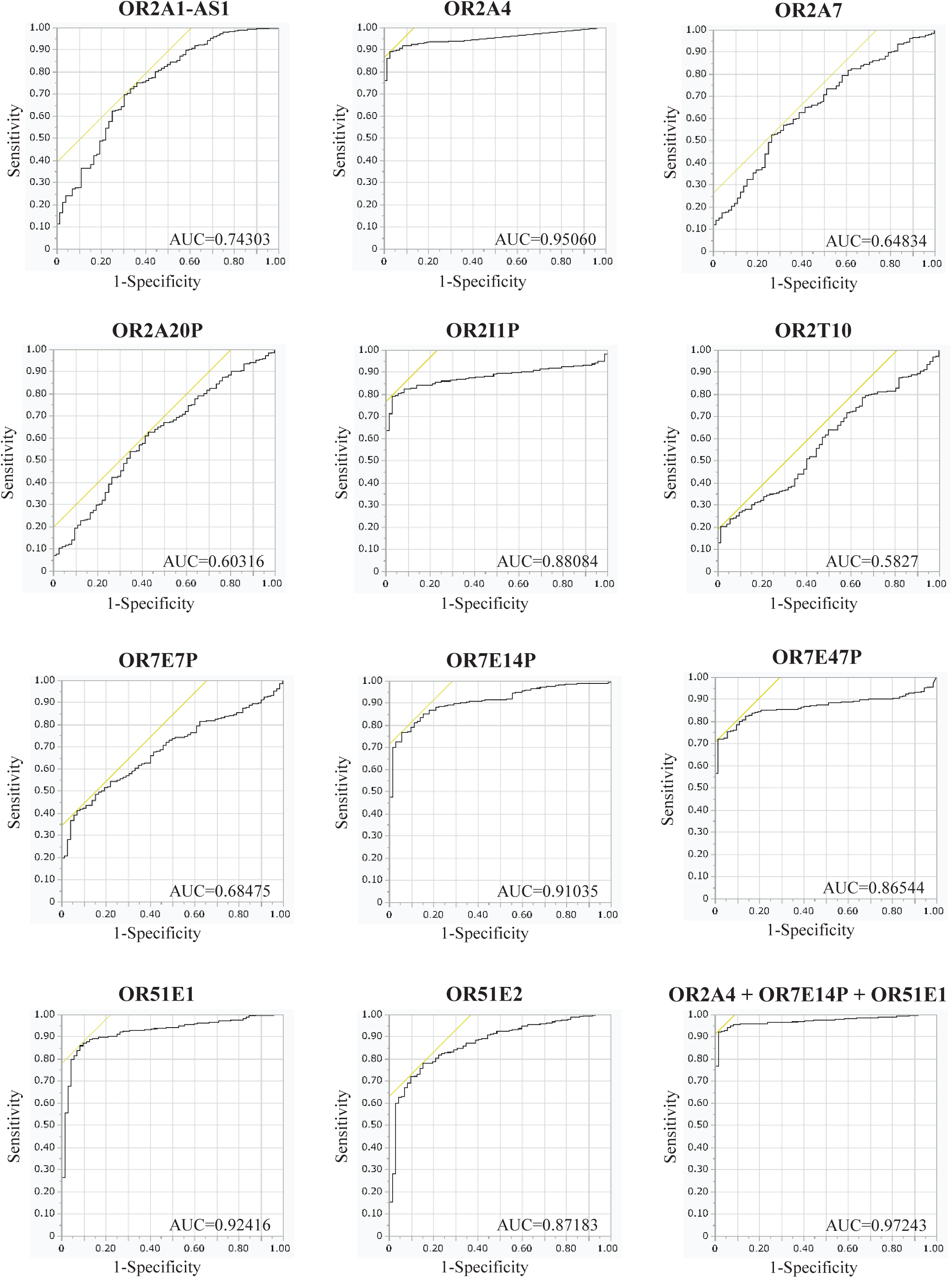
Receiver operating characteristic curve analysis of OR genes for the diagnosis of kidney cancer. ROC curves of OR2A4, OR51E1, and OR7E14P for discriminating between normal and tumor samples. The AUC values are shown.

### Association between OR Gene Expression and Patient Prognosis

To investigate whether OR gene expression is associated with the prognosis of patients with kidney cancer, we performed Kaplan-Meier analysis. Tumor samples were divided into high and low expression groups based on the median expression level of each OR gene. We found that high OR2A20P expression and low OR7E7P expression were significantly associated with poorer prognosis (Log-rank test, p = 0.0419 and p = 0.0155, respectively, Figure 6). Additionally, the Kaplan-Meier analysis suggested that high OR2A7 expression tended to be associated with poorer prognosis (p = 0.0710) (Figure 6).

**Figure 6.**
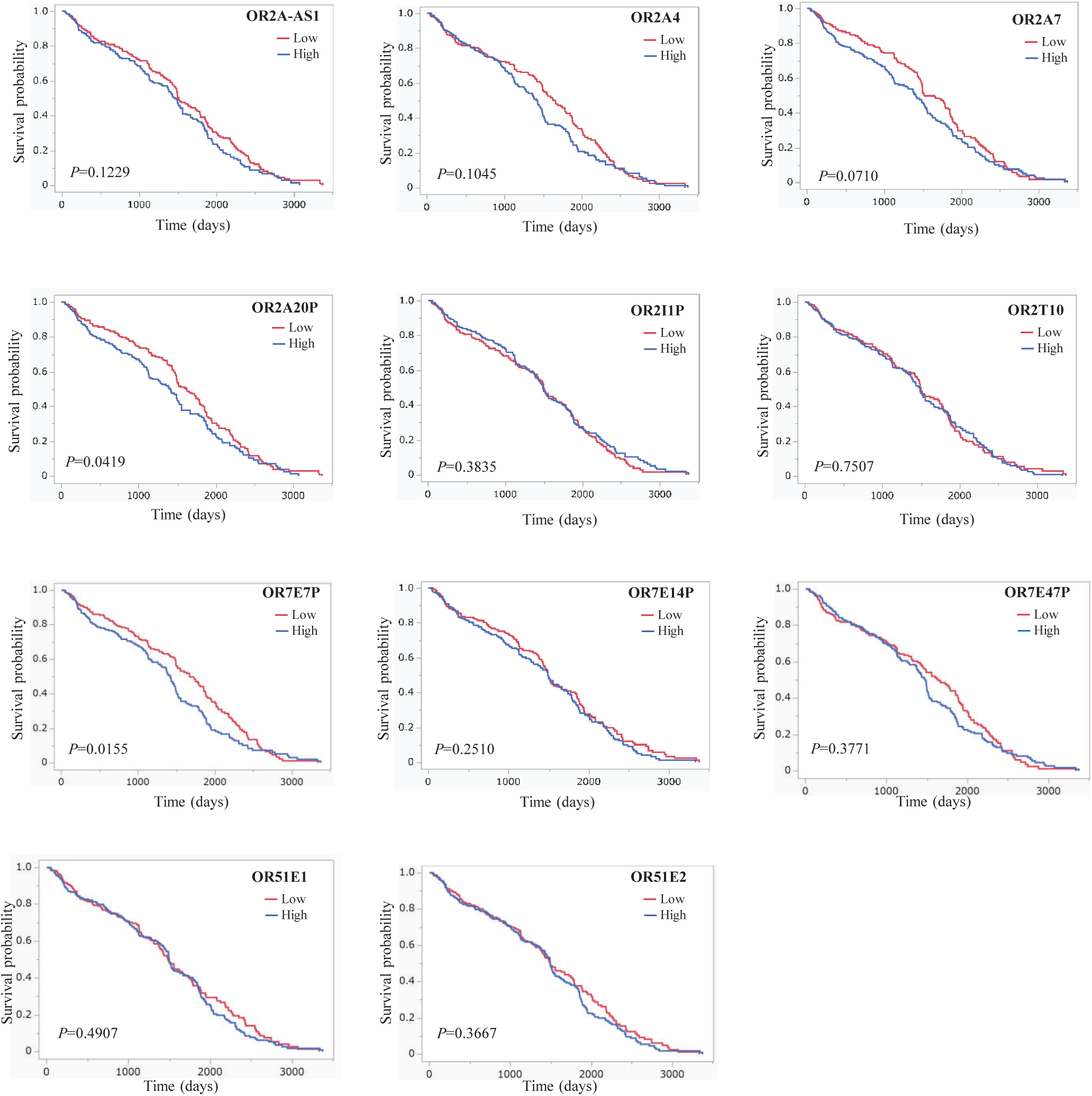
Kaplan-Meier survival analysis of kidney cancer patients according to OR gene expression. Kaplan-Meier curves for patients with high and low expression of OR2A20P, OR7E7P, OR2A7, OR2A1-AS1, OR2A4, OR2I1P, OR2T10, OR7E14P, OR7E47P, OR51E2, OR51E1. P-values were calculated using log-rank test.

### Sex-Based Differences in OR Gene Expression in Tumors

We examined whether there were sex-based differences in the expression of OR genes. We found significant differences in the expression of three genes (OR2A7, OR2I1P, and OR7E14P) between male and female patients (Figure 7). Specifically, these three genes were all significantly more strongly expressed in female patients.

**Figure 7.**
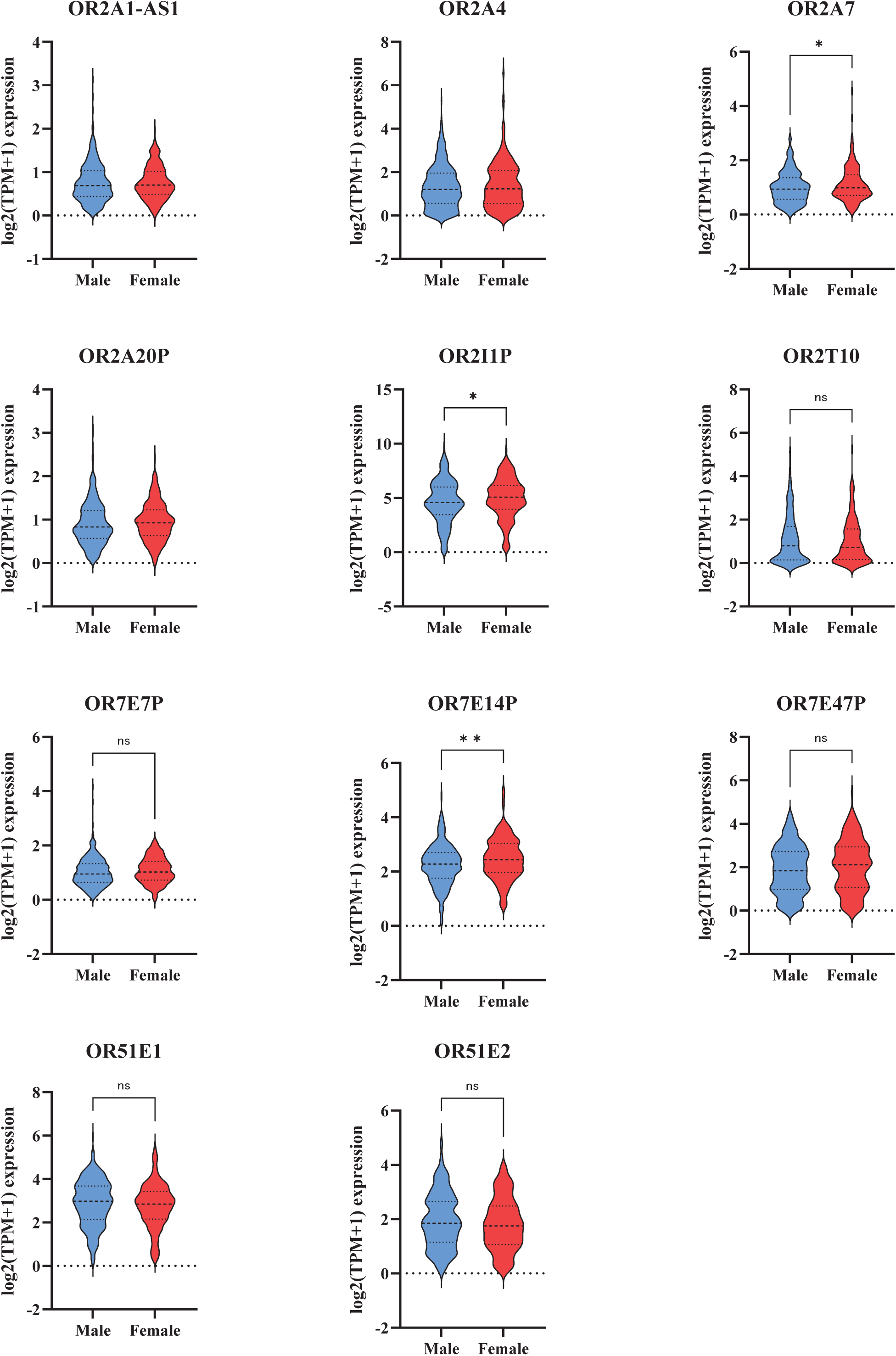
Sex-based differences in OR gene expression in the tumor group. Box plots showing the expression levels of OR2A7, OR2I1P, and OR7E14P in male (n=340) and female (n=187) tumor samples. *p < 0.05, **p < 0.01 (Mann-Whitney U test).

## Discussion

In this study, we comprehensively analyzed the expression and clinical significance of olfactory receptor (OR) genes in clear cell renal cell carcinoma (KIRC) using the TCGA KIRC dataset. Previous studies have reported the involvement of various ORs in cancer development and progression [20]. For example, Sanz et al. demonstrated that OR1A2 activation promotes cancer cell invasiveness and metastasis [20]. Our results revealed that the expression of multiple OR genes was significantly altered in KIRC. Notably, OR2A4, OR51E1, and OR7E14P emerged as potential diagnostic markers, whereas OR2A20P and OR7E7P showed promise as prognostic markers. Furthermore, we identified sex-based differences in the expression of OR2A7, OR2I1P, and OR7E14P, suggesting their potential in personalized medicine.

Comparing the normal and tumor groups, we found that 10 OR genes exhibited significant expression changes. Specifically, six genes (OR2A4, OR51E1, OR51E2, OR7E47P, OR7E7P, and OR2I1P) were upregulated in the tumor group and four genes (OR2A1-AS1, OR2A7, OR7E14P, and OR2A20P) were downregulated. In KIRC, clear cell RCC (ccRCC) is the most common histological subtype of renal cell carcinoma (RCC), and the TCGA KIRC dataset analyzed in this study mainly consisted of ccRCC cases. These genes may be involved in the development and progression of KIRC, particularly ccRCC. Specifically, OR genes upregulated in the tumor group may promote tumor cell proliferation and survival, whereas those downregulated may act as tumor suppressors.

These genes could be important molecular targets for understanding the pathogenesis of KIRC. Further functional analyses of these genes using in vitro and in vivo experimental systems are expected to shed light on the mechanisms of KIRC development and progression.

ROC analysis revealed that OR2A4, OR51E1, and OR7E14P exhibited high AUC values (0.951, 0.924, and 0.910, respectively) even when considered alone, indicating excellent diagnostic performance. Moreover, combining these three genes resulted in an AUC of 0.972, demonstrating extremely high accuracy in discriminating KIRC from normal tissues. These findings strongly suggest that OR2A4, OR51E1, and OR7E14P are useful diagnostic markers for KIRC. In particular, the combination of these three genes improved diagnostic performance compared with that of individual markers, raising expectations for clinical application. The combination of these genes may enable highly accurate detection of KIRC, and future studies should focus on developing diagnostic kits using these genes and constructing diagnostic systems in combination with other markers. Liquid biopsy approaches, such as the detection of circulating tumor cells or cell-free DNA in blood, can also be employed to detect these OR genes in a minimally invasive manner [21].

The analysis according to tumor stage (Stage) showed significant differences in the expression of OR2I1P, OR51E1, and OR7E14P, but no clear stage-dependent expression changes were observed. In other words, these genes did not exhibit a gradual increase or decrease in expression from Stage I to Stage IV. This suggests that many of the OR genes identified in this study may undergo expression changes from the early stages of tumorigenesis, potentially contributing to tumor initiation and progression. However, due to the limited sample size, further validation using a larger dataset is necessary.

Here, we outline the findings reported in previous studies on the OR genes that were particularly present in this study. OR51E1 is highly expressed in prostate cancer cells and promotes cell proliferation and resistance to apoptosis [22]. Furthermore, OR51E1 has been reported to be upregulated in other cancers, such as lung cancer [23], colon cancer [24], bladder cancer [23], and gastric cancer [25], suggesting its involvement in the malignancy of multiple cancers. In this study, the high expression of OR51E1 in the KIRC group suggests that this gene may also be involved in the development and progression of kidney cancer, which is consistent with the findings in prostate cancer and other cancers. The activation of OR51E1 promotes cancer cell proliferation and metastasis through the PI3K/Akt signaling pathway [24]. Concerning OR2A4, its expression is regulated via the androgen receptor in prostate cancer cells and is involved in cell proliferation [17]. In KIRC, OR2A4 may also promote the proliferation of tumor cells, and further studies, including a relationship with androgen receptor signaling, are anticipated. OR2A7 is expressed in an androgen-dependent manner in prostate cancer cells and is involved in cell proliferation [17]. In this study, OR2A7 was underexpressed in the KIRC tumor group but was expressed more in men than in women, suggesting a link with androgen signaling. Meanwhile, OR7E7P was reported to act as a growth inhibitor in breast cancer cells [26]. In KIRC, OR7E7P may have a similar tumor-suppressive function, and future functional analyses are warranted. Although the functional role of OR7E14P in cancer has not been extensively reported, its expression is reportedly regulated by the estrogen receptor in breast cancer cells [26]. In this study, OR7E14P was underexpressed in the KIRC tumor group and overexpressed in women, suggesting a link with estrogen signaling. Further research is needed to clarify the function and expression control mechanisms of OR7E14P, including in gynecological cancers other than breast cancer. OR2A20P and OR2I1P have been reported to exhibit altered expression in various cancers [27], but their roles in KIRC have not been reported. In this study, high OR2A20P expression was associated with poor prognosis in patients with KIRC, and OR2I1P was overexpressed in women. Although these genes have not been investigated in previous cancer research, the results of this study suggest that they may play an important role in the diagnosis, prognosis, and sex-specific personalized medicine of KIRC.

Among the OR genes that showed high expression in the KIRC tumor group in this study, OR51E1 and OR51E2 have been reported to be upregulated in other cancers, such as prostate cancer [22], lung cancer [23], colon cancer [24], bladder cancer [23], and gastric cancer [25]. These results suggest that OR51E1 and OR51E2 are commonly involved in the malignancy of multiple cancers. On the other hand, among the OR genes that showed low expression in the KIRC tumor group (OR2A1-AS1, OR2A7, OR7E14P, and OR2A20P), downregulation of OR2A1-AS1, OR2A7, and OR7E14P has not been reported in other cancers. Further studies are required to clarify whether the downregulation of these genes is a KIRC-specific phenomenon or a trend common to other cancers. Kaplan-Meier analysis revealed that high OR2A20P expression was significantly associated with poor prognosis, whereas high OR7E7P expression was significantly associated with good prognosis. Furthermore, high OR2A7 expression tended to be associated with poor prognosis (p=0.0710). These results suggest that OR2A20P, OR7E7P, and OR2A7 are useful prognostic markers in patients with KIRC. In particular, OR2A20P may be involved in tumor progression, and OR7E7P may have tumor-suppressive functions. Thus, examining the expression levels of these genes may provide information useful for predicting patient prognosis and determining treatment strategies. By combining the expression levels of these genes with clinicopathological factors, such as tumor stage and grade, more accurate prognostic predictions can be obtained. The identification of prognostic biomarkers is crucial for risk stratification and treatment selection in patients with KIRC [28].

Interestingly, we observed sex-based differences in the expression of OR genes in the tumor group. Specifically, OR2A7, OR2I1P, and OR7E14P were all significantly more strongly expressed in females. This result suggests that sex-related factors, such as sex hormones, are involved in the development and progression of KIRC. It is also possible that male-female biological differences affect the regulation of OR gene expression. Previous studies have reported an association between the expression of sex hormone receptors and OR genes in breast cancer [26], and a similar association may exist in KIRC. OR gene expression analysis may be useful for developing personalized medicine that considers sex differences. In particular, the development of therapeutic methods targeting the expression of these genes in female patients with KIRC is anticipated. Sex-based differences in cancer incidence, prognosis, and treatment response have been increasingly recognized, highlighting the need for sex-specific approaches in cancer management [29].

The limitations of this study include its retrospective nature based on TCGA data, bias in sample size (especially the small number of normal samples), and unknown functional roles of the OR genes. The small number of normal samples may have affected the interpretation of the results. Future studies should include analyses using a larger number of normal samples and prospective validation. Furthermore, although this study analyzed the association between OR gene expression and clinical information, the functional roles of OR genes require further investigation. For example, it is necessary to elucidate the signaling pathways through which ORs exert their effects on cancer cells. Recent studies have shown that ORs can activate various intracellular signaling cascades, including the cAMP/PKA and MAPK pathways [18, 19]. It is also important to investigate the mechanisms that regulate OR gene expression in cancer cells. Epigenetic modifications, such as DNA methylation and histone modification, play crucial roles in the regulation of gene expression in cancer [30].

Future prospects include validation of the results in independent large-scale cohorts, evaluation of clinical usefulness in prospective studies, functional analysis through in vitro and in vivo experiments, elucidation of the regulatory mechanisms of OR gene expression, and development of new therapies targeting ORs. The use of advanced gene editing technologies, such as CRISPR-Cas9, may facilitate the functional characterization of ORs in KIRC [31]. In particular, it is important to clarify the role of OR genes in the development and progression of KIRC through functional analysis in cellular and animal models. Further analysis of epigenetic control mechanisms, such as the methylation of the promoter regions of OR genes and transcription factors that regulate the expression of OR genes, is also an important research area in the future. In addition, the development of novel therapeutic strategies targeting ORs, such as small-molecule inhibitors and antibody-based therapies, is anticipated. ORs belong to the GPCR superfamily and are therefore considered promising drug targets [32]. Future studies should also consider using established cell lines, such as those described by Kato et al., to further investigate the functional roles of ORs in KIRC [33].

In conclusion, this study elucidated the expression profiles of OR genes in KIRC and demonstrated their potential as diagnostic, prognostic, and therapeutic markers. In particular, OR2A4, OR51E1, and OR7E14P are promising diagnostic markers, and OR2A20P and OR7E7P are promising prognostic markers. Their clinical application is anticipated in the future. In addition, the existence of sex differences in the expression of OR genes is an important finding for the realization of personalized medicine. The results of this study are expected to contribute to the early detection of KIRC, prediction of prognosis, and realization of personalized medicine, as well as the identification of new therapeutic targets.

## Data Availability

All data produced in the present study are available upon reasonable request to the authors

## Author Approvals

All authors have read and approved the final manuscript. The manuscript has not been accepted or published elsewhere.

## Acknowledgment

This work was supported by JSPS KAKENHI Grant Numbers JP23K27975 and JP24K22277, and by Takahashi Industrial and Economic Research Foundation Grant Number 112 (2024).

## References

1. Sung H, Ferlay J, Siegel RL, Laversanne M, Soerjomataram I, Jemal A, Bray F. Global Cancer Statistics 2020: GLOBOCAN Estimates of Incidence and Mortality Worldwide for 36 Cancers in 185 Countries. CA Cancer J Clin. 2021 May;71(3):209–249.

2. Xia, C., Dong, X., Li, H., et al. Cancer statistics in China and United States, 2024: profiles, trends, and determinants. Chin Med J (Engl). 2024;137(1):37–53.

3. An JY, Kim JH, Kim YJ, Oh SJ, Kim YK. Therapeutic Strategies for the Treatment of Renal Cell Carcinoma: An Update. Int J Mol Sci. 2023;24(21):15669.

4. Hsieh JJ, Purdue MP, Signoretti S, Swanton C, Albiges L, Schmidinger M et al. Renal cell carcinoma. Nat Rev Dis Primers. 2017;3:17009.

5. Linehan, W.M., Ricketts, C.J. The Cancer Genome Atlas of renal cell carcinoma: findings and clinical implications. Nat Rev Urol. 2019 Nov;16(11):539–552.

6. Wang X, Zhu X, Zhang Q, Li Y, Li P, Yang L, et al. Comprehensive investigation of olfactory receptor family for cancer diagnosis, prognosis, and immunotherapy. Front Immunol. 2024;15:1334404.

7. Capitanio U, Bensalah K, Bex A, Boorjian SA, Bray F, Coleman J et al. Epidemiology of Renal Cell Carcinoma. Eur Urol. 2016 Jan;69(1):32–45.

8. Rini BI, Campbell SC, Escudier B. Renal cell carcinoma. Lancet. 2009;373(9669):1119–1132.

9. The Cancer Genome Atlas Research Network. Comprehensive molecular characterization of clear cell renal cell carcinoma. Nature. 2013 Jul 4;499(7456):43–9.

10. Niimura Y, Nei M. Extensive gains and losses of olfactory receptor genes in mammalian evolution. PLoS One. 2007;2(8):e708.

11. Siegel RL, Miller KD, Jemal A. Cancer statistics, 2019. CA Cancer J Clin. 2019 Jan;69(1):7–34.

12. Heng DY, Albiges L. Progress in the systemic treatment of advanced renal cell carcinoma. Eur Urol. 2020;78(4):526–536.

13. Barata PC, Rini BI. Treatment of renal cell carcinoma: Current status and future directions. CA Cancer J Clin. 2017;67(6):507–524.

14. Nebert DW. “Olfactory” receptors: a misnomer. Genomics. 2017 Jul;109(3-4):125–133.

15. Feldmesser E, Olender T, Khen M, Yanai I, Lancet D, Ben-Asher E. Widespread ectopic expression of olfactory receptor genes. BMC Genomics. 2006;7:121.

16. Zhang X, De la Cruz O, Pinto JM, Nicolae D, Firestein S, Gilad Y. Characterizing the expression of the human olfactory receptor gene family using a novel DNA microarray. Genome Biol. 2007;8(5):R86.

17. Neuhaus EM, Zhang W, Gelis L, Deng Y, Noldus J, Hatt H. Activation of an olfactory receptor inhibits proliferation of prostate cancer cells. J Biol Chem. 2009;284(24):16218–16225.

18. Busse, D., Kudella, P., Grüning, N. M., et al. A synthetic sandalwood odorant induces wound healing in human keratinocytes via the olfactory receptor OR2AT4. J Invest Dermatol. 2014;134(11):2823–2832.

19. Massberg S, Simon MI. Phorbol ester-induced activation of the mitogen-activated protein kinase cascade in human neutrophils. Blood. 1999;93(8):2713–2720.

20. Sanz G, Leray I, Dewaele A et al. Promotion of cancer cell invasiveness and metastasis emergence caused by olfactory receptor OR1A2 activation. PLoS One. 2014;9(1):e85110.

21. Lin Y, Yang Z, Xu W et al. Cell-free DNA as a noninvasive biomarker for renal cell carcinoma: a systematic review and meta-analysis. J Cancer Res Clin Oncol. 2021;147(4):1005–1020.

22. Xu, L. H., Duan, X. H., Hu, Y., et al. Olfactory receptor 51E1 promotes proliferation and metastasis of prostate cancer cells via PI3K/Akt pathway. Oncol Rep. 2018;40(2):1094–1102.

23. Rodriguez M, Luo W, Weng J, Zeng L, Yi Z, Siwko S et al. An olfactory receptor as a potential target for the diagnosis and treatment of human transitional cell carcinoma of the bladder. Oncotarget. 2016;7(46):75998–76013.

24. Wu W, Shi D, Zhang J et al. Olfactory receptor 51E1 promotes tumor progression through the PI3K/AKT/mTOR signaling pathway and serves as a prognostic marker in colon cancer. Am J Cancer Res. 2020;10(10):3434–3449.

25. Giandomenico, V., Cui, T., Grimelius, L., et al. Olfactory receptor 51E1 expression is elevated in endocrine tumors of the pancreas and stomach. Oncotarget. 2013;4(1):44–53.

26. L. Weber, K. Al-Refae, S. Ebner, et al. Expression and functionality of olfactory receptors on human breast cancer cells. Front Oncol. 2018;8:33.

27. Cui T, Chen Y, Knösel . Whole-exome sequencing identifies recurrent somatic mutation in olfactory receptor genes in hepatocellular carcinoma. Medicine (Baltimore). 2016;95(46):e5337.

28. Zhang Z, Li Y, Ding Y et al. Development and validation of a prognostic nomogram for clear cell renal cell carcinoma based on immune-related genes. Front Immunol. 2021;12:632951.

29. Cooke VG, Freiman A, Mayo NE, et al. Sex, gender, and cancer: an update on progress and gaps. Lancet Oncol. 2023;24(11):e463–e475.

30. Jones PA, Baylin SB. The fundamental role of epigenetic events in cancer. Nat Rev Genet. 2002;3(6):415–428.

31. Hsu PD, Lander ES, Zhang F. Development and applications of CRISPR-Cas9 for genome engineering. Cell. 2014;157(6):1262–1278.

32. Overington JP, Al-Lazikani B, Hopkins AL. How many drug targets are there? Nat Rev Drug Discov. 2006;5(12):993–996.

33. Kato K, Ohkura M, Kuno T et al. Establishment of a cell line from a mouse xenograft model of clear cell renal cell carcinoma and its in vivo growth. Oncol Lett. 2019;18(4):3558–3564.

